# Clinical efficacy and safety assessment of specific model electroacupuncture stimulation in enhancing paclitaxel delivery across the blood-brain barrier for recurrent malignant glioma: study protocol of a single-arm trial

**DOI:** 10.1101/2025.08.01.25332715

**Authors:** Zhaoxing Jia, Tianxiang Jiang, Yiqing Zhang, Qianyue Chen, Zhong Di, Qi Yuan, Kecheng Qian, Lin Gan, Congcong Ma, Xianming Lin

## Abstract

**Introduction:** Despite advances in surgical techniques, radiotherapy, and chemotherapy, the prognosis for patients with recurrent malignant gliomas remains poor. Although paclitaxel is widely used for peripheral solid tumors and demonstrates potent antitumor effects in glioma cell lines in vitro, its clinical efficacy against recurrent malignant gliomas via intravenous administration remains suboptimal due to blood-brain barrier (BBB) restrictions. Recent studies have demonstrated that specific-mode electrical stimulation (SMES) can transiently open the BBB, thereby enhancing tumor accumulation of albumin-bound paclitaxel (ABX) and exerting anti-glioma effects. This study aims to evaluate the therapeutic efficacy and safety profile of SMES combined with ABX in patients with recurrent malignant gliomas.

**Methods and analysis:** This is a single-center, single-arm, prospective phase II clinical trial. A total of 20 patients with histologically confirmed recurrent malignant gliomas (WHO grade 4) will be enrolled. Eligible patients will receive intravenous ABX (135–175 mg/m²) every 21 days for a total of 6 cycles. A Simon’s two-stage design will be employed, with the primary endpoint being the 4-month progression-free survival (PFS) rate. Secondary endpoints include adverse events (AEs), disease control rate (DCR), objective response rate (ORR), duration of disease control (DDC), duration of response (DOR), Neurological Assessment in Neuro-Oncology (NANO) score, European Organisation for Research and Treatment of Cancer Quality of Life Questionnaire-Core 30 (EORTC QLQ-C30), progression-free survival (PFS), and overall survival (OS).

**Ethics and dissemination:** This study was approved by the Medical Ethics Committee of the Third Affiliated Hospital of Zhejiang Chinese Medical University. The study findings will be presented at international conferences and published in peer-reviewed journals.

**Trial registration number:** NCT06818331

**STRENGTHS AND LIMITATIONS OF THIS STUDY:** ⇒ This study explores potential therapeutic options for refractory diseases characterized by the absence of effective standard therapies and short survival expectancy.

⇒ A key strength of this trial lies in its ability to yield rapid results, thereby optimizing resource allocation.

⇒ Comparable clinical data demonstrate that the safety profile of this regimen is predictable, acceptable, and manageable.

⇒ The primary limitation involves the single-arm design, wherein comparison with external historical controls may introduce bias and compromise the strength of conclusions.

⇒ The modest sample size may result in insufficient statistical power for secondary endpoint analyses.

⇒ Despite standardized assessment protocols, the intrinsic heterogeneity of neuro-oncological malignancies may affect the consistency of therapeutic response evaluations.

## BACKGROUND

Glioblastoma multiforme (GBM), the most aggressive malignant neoplasm in the central nervous system, exhibits a global annual incidence of 3-5 cases per 100,000 population(^1^). Notably, GBM accounts for approximately 50% of all gliomas, representing the predominant histologic subtype(^2^).The majority of GBM cases present with insidious onset, typically reaching advanced stages at diagnosis(^3^). Merely 30% of patients qualify for radical surgical resection(^4^), and even with adjuvant chemoradiotherapy, the median PFS rarely exceeds 7 months(^5^).

Prognosis remains dismal, with standard therapy yielding a median overall survival of 12-15 months(^5, 6^) and near-universal recurrence rates approaching 100%(^7, 8^).

Paclitaxel, a microtubule-stabilizing agent, exerts antitumor effects by disrupting mitotic spindle dynamics and inducing apoptosis, rendering it a cornerstone therapy for diverse solid malignancies. In vitro studies demonstrate potent antitumor activity of paclitaxel in glioma cell lines (e.g., U87, U251), with mean IC50 values 1400-fold lower than temozolomide(^9–12^). However, clinical trials have revealed limited efficacy of intravenous paclitaxel monotherapy in recurrent glioblastoma(^13, 14^), primarily attributable to insufficient drug accumulation in tumor tissue due to blood-brain barrier exclusion(^15, 16^), thereby precluding therapeutic efficacy.

To overcome blood-brain barrier (BBB) limitations, various paclitaxel delivery strategies have been developed, including drug structural modifications and carrier-mediated delivery systems to enhance BBB penetration(^17–20^). Recent breakthroughs in focused ultrasound-induced BBB opening demonstrate 3-5-fold increases in cerebral paclitaxel concentration in animal models, with significant survival extension. The first human trial (NCT04528680, 2021) preliminarily confirmed the safety of this approach, showing local disease control in 42% of patients(^21, 22^).However, despite improved drug delivery to the brain, these strategies face critical challenges including inconsistent delivery efficiency, safety concerns, and high treatment costs, limiting clinical translation. Therefore, there is an urgent need for safer, more operable and cost-effective BBB-penetrating paclitaxel delivery strategies.

Our preliminary studies demonstrated that specific model electroacupuncture stimulation (SMES: 2/100Hz, 3mA, 6s-on/6s-off, 40min) effectively opens the BBB, facilitating macromolecule penetration including Evans blue and FITC-dextran(^23^). SMES-facilitated neural growth factor delivery across the blood-brain barrier significantly improved learning and memory functions in MCAO/R model rats during the recovery phase(^24^). The BBB-opening effect of SMES exhibits rapid on/off switching without inducing cerebral edema or neuroinflammation, confirming its safety and controllability(^25^). In glioma-bearing mice, SMES increased tumor accumulation of ABX through BBB modulation, resulting in anti-glioma effects(^26^).

Although our studies have demonstrated SMES-mediated macromolecular drug delivery across the BBB, enhancing intracerebral accumulation and therapeutic efficacy, prospective clinical trials are required to validate the clinical benefits of SMES-induced BBB opening for ABX delivery in malignant glioma treatment. Accordingly, we initiated a single-arm, single-center phase II clinical trial to evaluate the safety and efficacy of SMES-ABX combination therapy for malignant gliomas.

## METHODS

### Study design

This study is a single-center, single-arm, open-label phase II clinical trial to be conducted at the Third Affiliated Hospital of Zhejiang Chinese Medical University. The protocol was approved by the Institutional Review Board on January 15, 2025 (approval number: ZSLL-KY-2024-079-01) and has been registered on ClinicalTrials.gov (registration number: NCT06818331). Patient enrollment will commence in May 2025 with anticipated completion by January 2028. Eligible malignant glioma patients will receive SMES-ABX combination therapy, with 4-month progression-free survival (4m-PFS) rate serving as the primary endpoint. Secondary endpoints include: AEs, DCR, ORR, DDC, DOR, NANO scale, EORTC QLQ-C30, PFS, and OS.The study protocol adheres to the Standards for Reporting Interventions in Clinical Trials of Acupuncture (STRICTA 2010) and SPIRIT guidelines(^27, 28^).

### Participant selection

In this study, postoperative recurrence of malignant glioma is defined as meeting all criteria: (1) WHO grade IV classification per the Chinese Anti-Cancer Association Guidelines for Integrated Diagnosis and Treatment of Glioma (V2.0_2025, 20250110)(^29^); (2) prior surgical resection; (3) neuroradiologically confirmed recurrence via cranial MRI, including either progression of original lesions or emergence of new lesions, as adjudicated by board-certified neuro-oncologists. Subject selection will follow predefined inclusion/exclusion criteria.

### Inclusion criteria

- Histologically confirmed WHO grade IV glioma per Chinese Anti-Cancer Association Guidelines for Integrated Diagnosis and Treatment of Glioma (V2.0_2025, 20250110).
- Radiologically confirmed disease progression per RANO 2.0 criteria following surgical resection, with ≥1 measurable contrast-enhancing lesion.
- Age 18-70 years (inclusive), any gender.
- Dexamethasone dose for mass effect: <6mg daily (stable for 7 days) or <6mg average during tapering. Non-mass-effect steroid use permitted.
- Functional status: KPS ≥40.
- Adequate organ function (within 14 days):

a. Hemoglobin ≥90g/L
b. WBC ≥3.0×109/L
c. ANC ≥1500/µL (WBC×neutrophil %)
d. Platelets ≥100×109/µL
e. Total bilirubin ≤5×ULN
f. AST ≤3×ULN (with bilirubin ≤3×ULN)
g. Creatinine ≤1.5mg/dL or eGFR 30-90mL/min
- Tolerability of electroacupuncture with expected compliance.
- Conscious with preserved:

– Nociception/discrimination
– Basic communication capacity
- Willingness to provide written informed consent.

### Exclusion criteria

- Uncontrolled seizure disorder.
- Concurrent participation in other interventional trials or within 30 days post-participation.
- Currently receiving paclitaxel or similar drug treatment.
- Severe allergy to paclitaxel or similar compounds
- Pregnant or breastfeeding women.
- Diseases affecting cognitive function such as congenital dementia, or alcoholism, drug abuse or psychotropic substance abuse.
- Skin infection at acupuncture sites.
- Patients with conductive foreign objects in body.
- Contraindications to gadolinium-enhanced MRI.
- Other acute or chronic diseases, mental illnesses or abnormal laboratory test values that may increase risks associated with study participation or study drug administration, or interfere with interpretation of study results, and determined by investigator as ineligible for study participation.
- Concurrent other types of anti-tumor treatments during trial such as chemotherapy, radiotherapy, targeted therapy, immunotherapy etc..

### Baseline evaluation

Following screening and enrollment, baseline assessments will be conducted within 48 hours. Comprehensive data collection will include: (1) informed consent documentation, (2) medical history, (3) physical examination, (4) laboratory tests, (5) MRI scans (contrast-enhanced brain MRI, MR spectroscopy, perfusion-weighted imaging, etc.), (6) NANO scale, (7) EORTC QLQ-C30, (8) tumor characteristics (pathological and genomic analyses), and (9) detailed demographic information. Further specifications are provided in Fig 1.

**Fig 1.**
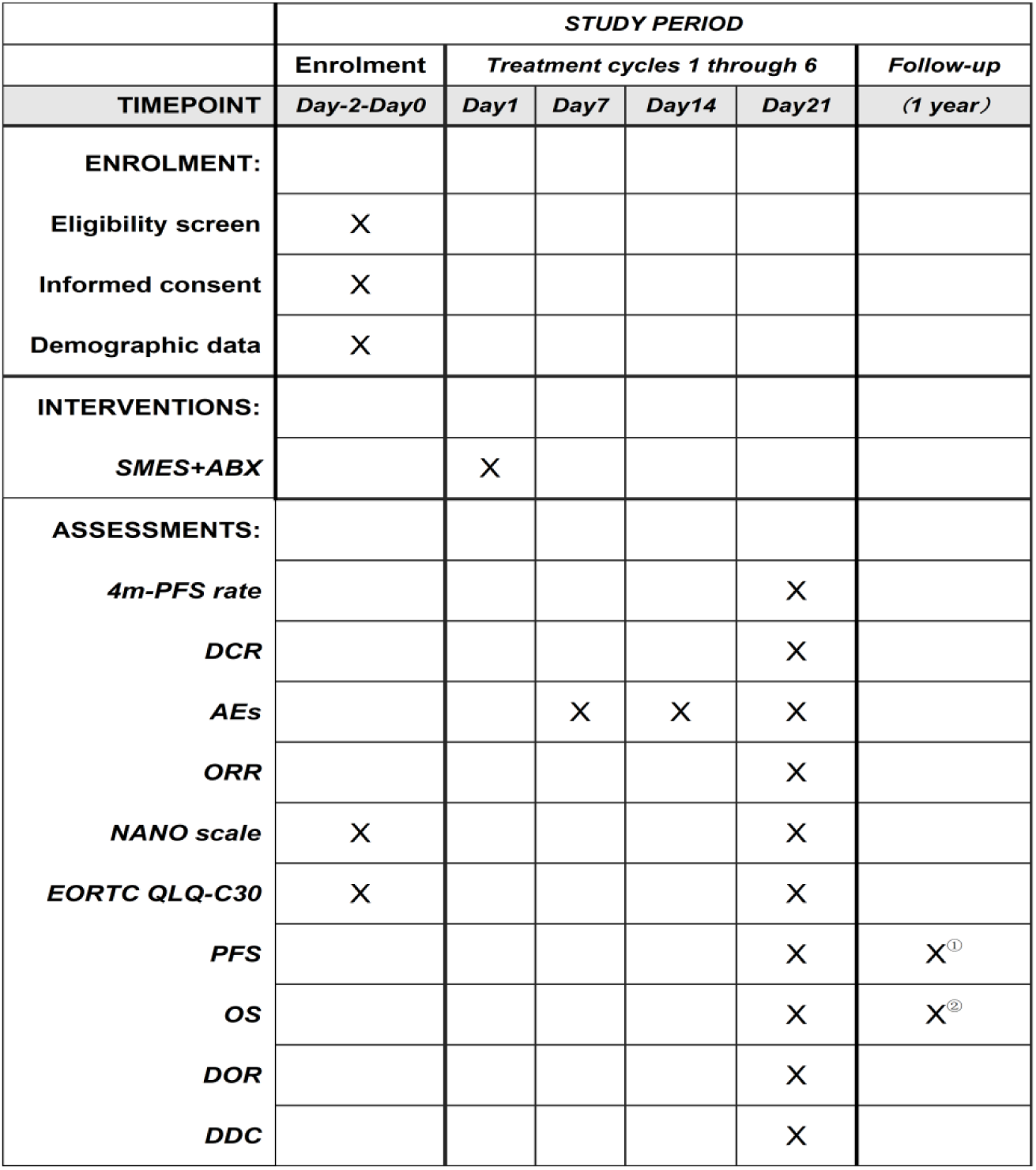
SPIRIT schedule of enrolment, interventions, and assessments. ①② Monthly follow-up assessments were conducted during the observation period. 4m-PFS rate; 4-month progression-free survival rate; DCR, disease control rate; AEs, adverse events; ORR, objective response rate; NANO, neurological assessment in neuro-oncology; EORTC QLQ-C30, european organisation for research and treatment of cancer quality of life questionnaire-core 30; PFS, progression-free survival; OS, overall survival;DOR, duration of response; DDC, duration of disease control.

### Interventional methods

Enrolled patients will receive intravenous ABX (135-175 mg/m²) on day 1 of each 21-day treatment cycle, with concurrent SMES intervention (Fig 2).

**Fig 2.**
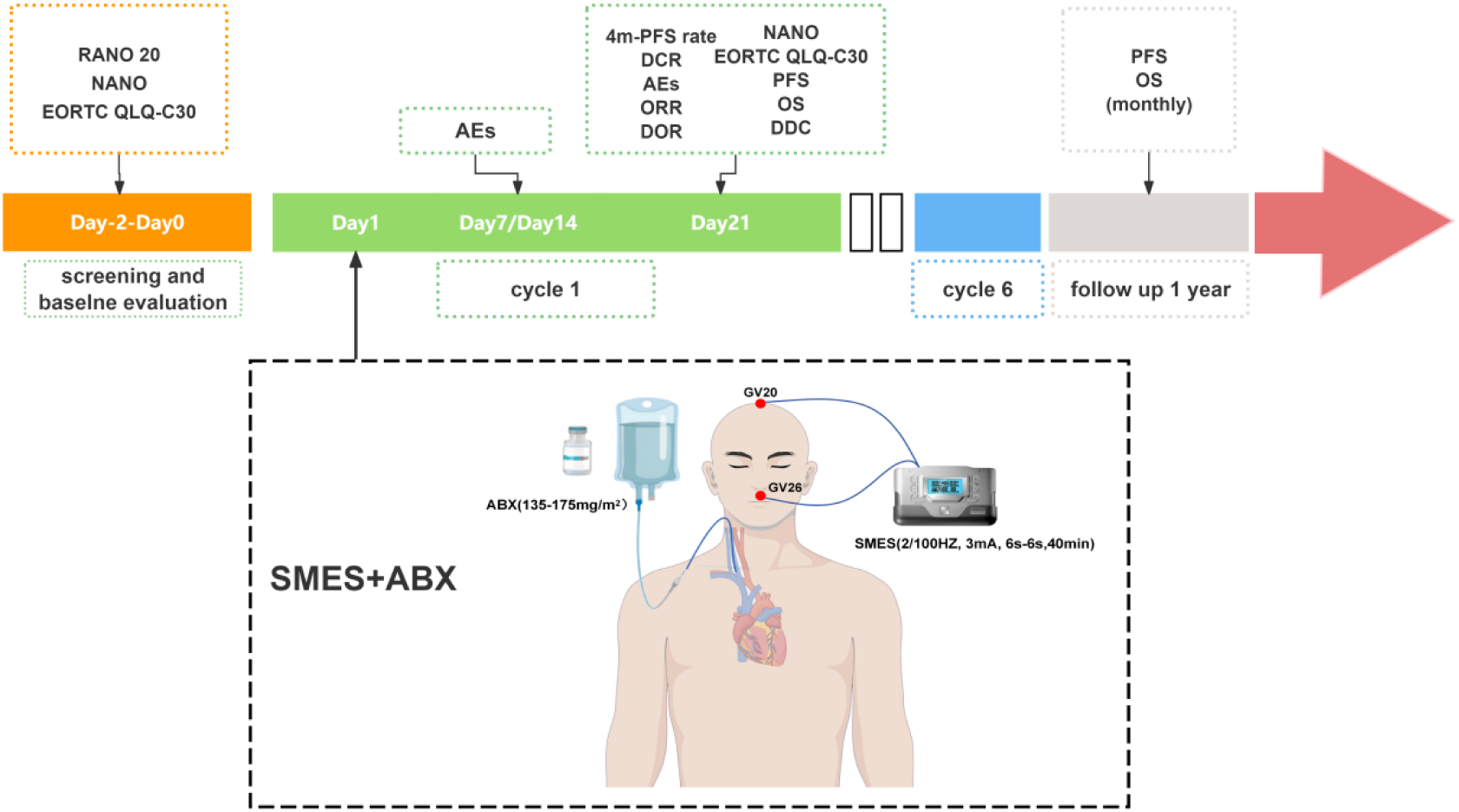
Schematic overview of the experimental protocol. The treatment regimen consisted of 6 cycles (21 days/cycle), with combined SMES and antibiotic (ABX) therapy administered on day 1 of each cycle. Efficacy assessments were performed on days 7, 14 and 21 of every cycle. Post-intervention monitoring continued for 12 months with monthly follow-up assessments.

### ABX treatment protocol

ABX was administered intravenously at 175 mg/m² over 40 minutes. Dose reduction to no less than 135 mg/m² was mandated for: (1) severe neutropenia (ANC <500/mm³ persisting ≥7 days), or (2) grade ≥3 sensory neuropathy. Treatment was withheld for grade 3 neurotoxicity until resolution to grade ≤2.

### SMES Therapeutic Protocol

The patient was placed in a supine position with full-body relaxation, followed by standard disinfection of the skin surrounding GV20 (Baihui) and GV26 (Shuigou). Acupuncture was then administered: at DU20, a 0.25mm × 40mm filiform needle was inserted horizontally backward (15–20 mm depth) using a twirling technique until local soreness and distension were achieved. At GV26, a 0.18mm × 25mm needle was obliquely inserted toward the nasal septum (9–15 mm depth) with lifting-thrusting and twirling manipulations until mild lacrimation or ocular moisture was observed. Subsequently, the needles were connected to an electroacupuncture device (HANS-200, Jisheng Medical Technology, Nanjing, China). Electrical stimulation was delivered in a dense-disperse wave mode (2/100 Hz) at 1.5–3.0 mA, adjusted to elicit mild muscle twitching near GV26. The treatment lasted 40 min, with a 6s ON/6s OFF duty cycle controlled by an external device.

### Assessment and follow-up

Tumor response will be evaluated according to the RANO 2.0 criteria(^30^), incorporating MRI findings and clinical outcomes to determine therapeutic efficacy. Baseline assessments will be conducted within 3 days prior to treatment initiation, followed by serial evaluations at 3-week intervals for a total of 7 assessments. AEs will be graded per CTCAE v5.0. Additionally, the causality between AEs and the investigational drug will be assessed to identify treatment-related adverse events (TRAEs). Furthermore, patient-reported outcomes, including quality of life and performance status, will be systematically evaluated.

Post-study follow-up will be conducted for 1 year, with monthly telemedicine consultations (via telephone or WeChat) to document survival outcomes. As illustrated in Fig 2, a comprehensive analysis will be performed for each parameter, with predefined time frames.

### Concomitant medications

Concomitant medications for AEs management were permitted during the trial, including: (1) acid-suppressing agents for gastric protection, (2) hepatoprotective drugs for liver function abnormalities, (3) low-dose corticosteroids for allergy prophylaxis, (4) recombinant human granulocyte colony-stimulating factor (rhG-CSF) or pegylated G-CSF to stimulate leukopoiesis, and (5) recombinant thrombopoietin (rhTPO) to promote thrombopoiesis. Detailed records were maintained for all concomitant medications, including: generic name, indication, single dose (with units), administration frequency, route, start date, and current status (ongoing/discontinued).

### Rationale for ABX Dose Selection

Although the manufacturer-recommended maximum dose of ABX for breast cancer is 260 mg/m², no established clinical guidelines exist for its use in malignant gliomas. Previous studies have demonstrated that ultrasound-mediated BBB disruption significantly enhances paclitaxel delivery in glioma models, achieving 3-5-fold higher brain concentrations compared to controls(^31^). Similarly, our findings indicate that SMES (sonication-mediated enhanced delivery) transiently disrupts the BBB, resulting in a 5.5-fold increase in ABX tumor accumulation relative to controls(^26^). Notably, a phase I clinical trial confirmed the safety of 260 mg/m² ABX when combined with repeated ultrasound-induced BBB opening(^21^).

However, our preliminary data revealed that SMES combined with ABX doses exceeding 175 mg/m² induced severe dose-limiting toxicities, primarily myelosuppression, resulting in significantly reduced treatment tolerability. Given the discrepancy between preclinical evidence and early clinical safety data, we selected a reduced ABX dose range (135–175 mg/m²) to ensure patient safety while maintaining therapeutic feasibility.The dosing strategy was determined by integrating the enhanced drug exposure resulting from blood-brain barrier disruption technology with a rigorous assessment of clinical tolerability, thereby ensuring both scientific validity and clinical relevance.

### Outcome measure primary endpoint

The primary objective of this study was to determine the 4-month progression-free survival (4m-PFS) rate, defined as the proportion of subjects in the intention-to-treat (ITT) population who remained progression-free beyond 4 months. This endpoint was continuously assessed throughout the study duration.

### Secondary endpoints

The DCR was defined as the proportion of patients in the ITT population who achieved CR, pPR), or stable disease (SD), as assessed per RANO 2.0 criteria.

The ORR was calculated as the percentage of ITT patients exhibiting CR or PR, evaluated according to RANO 2.0 guidelines.

The OS was measured from the initiation of SMES+ABX therapy until death or censoring at the last follow-up.

The PFS was defined as the time from first SMES+ABX administration to disease progression (per RANO 2.0), intolerable AEs, or death, whichever occurred first.

DOR was defined as the duration between the first SMES + ABX treatment and the occurrence of PD.

The DDC represented the time from initial achievement of disease control (CR/PR/SD) to PD or death.

The NANO scale is a standardized quantitative instrument that evaluates neurofunctional status in neuro-oncology patients through systematic assessment of 8 key domains (e.g., motor, sensory, and language functions)

The EORTC QLQ-C30 questionnaire is a globally validated multidimensional instrument assessing cancer-related quality of life across five functional domains (physical, role, cognitive, emotional, and social) and symptom scales.

Safety assessments included treatment-emergent adverse events (TEAEs), treatment-related adverse events (TRAEs), and serious adverse events (SAEs). TEAEs were defined as any adverse events occurring between treatment initiation and study completion. TRAEs referred to adverse events specifically attributable to the investigational drug. SAEs were classified as events resulting in death, life-threatening illness, disability, hospitalization or prolonged hospitalization, or persistent/severe incapacity.

### Statistical analysis

#### Sample size

The sample size was determined using Simon’s two-stage design, with the primary efficacy endpoint being the 4m-PFS rate. Previous studies demonstrated that temozolomide treatment yielded a 4-m PFS rate no greater than 25.7% in patients with recurrent WHO grade 4 glioblastoma(^32, 33^). However, our preliminary data suggested that the SMES+ABX regimen could achieve a 4-m PFS rate of 60% in this patient population. Based on R 4.3.1 (clinfun package) calculations (one-sided α =0.05, power=80%), a total of 20 patients were planned, accounting for a 20% dropout rate. In the first stage, 9 patients were enrolled, with trial termination if fewer than 3 achieved 4-m PFS. Subsequently, an additional 7 patients were recruited in the second stage. The study would be deemed ineffective if the overall 4-m PFS rate was <56.3% (9/16 patients).

#### Statistical analysis methods

To comprehensively evaluate treatment efficacy and safety, four distinct analysis populations were predefined: 1) Intent-to-treat (ITT) population: all subjects who provided informed consent (regardless of actual treatment receipt); 2) Full analysis set (FAS): derived from the ITT population by excluding only clearly invalid data (e.g., immediate post-enrollment consent withdrawal); 3) Per-protocol set (PPS): a subset of FAS comprising subjects who strictly adhered to the study protocol; 4) Safety set (SS): a set of patients who had received at least one SMES + ABX treatment after enrollment and had a safety record.

A predefined analytical framework based on the RANO 2.0 criteria was employed. Time-to-event endpoints (including PFS, OS, DOR, and DDC) were analyzed using Kaplan-Meier survival curves, with between-group comparisons performed via log-rank tests. Additionally, multivariate analysis was conducted using Cox proportional hazards models. Categorical variables (e.g., ORR, DCR) were expressed as frequencies (percentages), and intergroup differences were assessed using Fisher’s exact test or χ² test, depending on sample size. Continuous variables (e.g., NANO score, QLQ-C30 scale) were analyzed for longitudinal changes using mixed linear models, while baseline-adjusted analysis of covariance (ANCOVA) was applied for cross-group comparisons. Safety data were graded per CTCAE v5.0 criteria, with adverse event incidence rates and 95% confidence intervals calculated using the Clopper-Pearson method. All tests were two-sided with α =0.05, and multiple comparisons were adjusted via false discovery rate (FDR) correction. Statistical analyses were performed using R 4.3.1 (survival and lme4 packages) and SAS 9.4. Sensitivity analyses, including bootstrap resampling (1000 iterations), were conducted to validate the robustness of key findings.

### Ethical Approval and Study Registration

This study received ethical approval from the Institutional Review Board of the Third Affiliated Hospital of Zhejiang Chinese Medical University (Approval No. ZSLL-KY-2024-079-01). Prior to enrollment, potential participants were comprehensively informed of the study protocol, anticipated benefits, and potential risks through standardized procedures. Written informed consent was obtained after confirming full comprehension. The trial strictly adhered to the principles of the Declaration of Helsinki. All research data were de-identified and stored in encrypted databases to ensure participant confidentiality. The study protocol was registered at ClinicalTrials.gov (Identifier: NCT06818331) and underwent continuous quality monitoring by an independent data monitoring committee.

### Data Monitoring

A standardized data management protocol was rigorously implemented throughout the study. Certified clinical research coordinators prospectively entered all study data into electronic case report forms (eCRFs) in compliance with the data management plan, ensuring both timeliness and accuracy. A three-tier quality control system was established, comprising: (1) source data verification, (2) logical consistency checks, and (3) medical monitoring, thereby safeguarding data authenticity, completeness, and reliability. All research data were stored in a centralized database with dual-layer encryption while maintaining comprehensive audit trails, thus providing a robust foundation for subsequent electronic data capture and analysis.

## DISCUSSION

### Therapeutic Efficacy of Paclitaxel in Malignant Glioma

Malignant gliomas, primarily encompassing WHO grade III and IV gliomas, represent a class of highly aggressive brain tumors with particularly poor prognosis(^34^). Current therapeutic modalities include surgical resection, radiotherapy, chemotherapy, targeted molecular therapy, tumor-treating fields, and oncolytic virotherapy, among others(^35, 36^). Despite multimodal therapeutic interventions, patient survival rates remain dismal, with multifocal high-grade gliomas demonstrating a median overall survival of merely 8 months(^37^).

Paclitaxel, a microtubule-stabilizing agent, is widely employed in chemotherapy for various malignancies including breast and ovarian cancers. Its antitumor mechanism involves stabilizing microtubules and suppressing their dynamics, thereby inducing mitotic arrest and subsequent apoptosis in proliferating cells(^38^). Glioblastoma demonstrates comparable susceptibility to paclitaxel as breast cancer(^39^). Data from the Cancer Cell Line Encyclopedia reveal potent antiglioma activity of paclitaxel in vitro, with an average IC50 approximately 1,400-fold lower than temozolomide(^40^). Preclinical studies have demonstrated paclitaxel’s superior efficacy over temozolomide in tumor growth suppression and survival prolongation in glioma-bearing rodent models(^41–43^). However, the clinically conventional ABX exhibits poor BBB penetration, failing to achieve therapeutic concentrations in brain tissue. Consequently, research efforts have focused on enhancing BBB penetration through three strategies: (1) nanomaterial-based formulation modification(^44^), (2) combination with BBB-modulating agents(^45^), and (3) physical/chemical BBB disruption(^46^). Nevertheless, these approaches face clinical translation challenges due to technical complexity and high costs, highlighting the urgent need for developing safe and effective strategies to deliver paclitaxel across the BBB while maintaining therapeutic concentrations in tumor tissue.

### Strengths of the study

In this study, SMES represents an innovative approach for BBB modulation. The intervention involved acupuncture at GV20 (Baihui) and GV26 (Shuigou) points, followed by connection to an electroacupuncture device (HANS-200, Jisheng Medical Technology, Nanjing, China) generating specific stimulation parameters (2/100Hz alternating frequency, 3mA intensity, 6s on/6s off duty cycle, 40min duration). SMES enhanced cerebral perfusion while reducing tight junction protein expression (ZO-1 and occludin) and inducing ultrastructural alterations in tight junctions(^25, 47^), resulting in reversible BBB permeability enhancement that resolved within 12 hours post-intervention. Importantly, this modality demonstrated an excellent safety profile, with no observed cerebral edema, glial activation, or apoptosis(^23^). A systematic review and meta-analysis confirmed that dense-disperse wave electroacupuncture enhances BBB penetration of Evans blue, potentially mediated through VEGF and MMP-9 upregulation(^48^). Our team further demonstrated that SMES facilitated BBB opening in glioma-bearing mice, significantly increasing tumor accumulation of albumin-bound paclitaxel and enhancing antitumor efficacy(^26^). Supporting this approach, a phase I clinical trial by Sonabend et al. established the safety of ultrasound-mediated BBB opening for ABX (260mg/m^2^) delivery in recurrent glioblastoma patients(^21^). However, the ultrasound approach requires surgical transducer implantation with associated technical complexity, high costs, and procedural risks, whereas SMES offers a non-invasive, cost-effective, and readily controllable alternative for clinical translation.

Secondly, the design of acupoint prescriptions. Two key acupoints, GV20 and GV26, were selected based on their established therapeutic relevance in neurological disorders. GV20 is anatomically localized along the midline of the head, 5 cun superior to the anterior hairline or at the midpoint between the bilateral auricular apices. DU26 is situated at the junction of the upper and middle thirds of the philtrum. According to Traditional Chinese Medicine (TCM) meridian theory, these acupoints are pivotal in managing cerebral pathologies due to their regulatory effects on neural-vascular pathways. Contemporary studies demonstrate that acupuncture at GV20 and GV26 stimulates the sphenopalatine ganglion and the trigemino-cerebrovascular system, inducing vasodilation of facial cerebral vasculature and significantly modulating hemodynamics (^49–53^). Notably, these interventions enhance regional cerebral blood flow velocity in specific brain areas.

Collectively, specific model electroacupuncture stimulation at GV20 and GV26 demonstrates potential for transient BBB disruption and enhanced paclitaxel accumulation in tumor tissues, as evidenced by our preliminary data. This study aims to evaluate the efficacy and safety of SMES combined with ABX therapy in patients with recurrent malignant glioblastoma (GBM), which may provide a novel therapeutic strategy for this refractory condition

### Limitations

Several methodological limitations of this study warrant explicit discussion. First, although the single-arm trial design was justified by the absence of standard therapies for recurrent malignant glioma, efficacy evaluation via historical controls may introduce selection bias, particularly due to inter-institutional variations in clinical management. Second, although the Simon’s two-stage design optimally controls type I error and enhances study efficiency, the limited sample size (N=16) may result in insufficient statistical power for secondary endpoint analyses. Notably, inherent heterogeneity in neuro-oncological manifestations may affect response assessment consistency, despite standardized RANO 2.0 criteria implementation. Nevertheless, as phase II trials are inherently exploratory rather than confirmatory, this study primarily aims to provide preliminary evidence for the SMES+ABX regimen’s potential superiority, with positive results warranting validation through subsequent phase III randomized controlled trials.

## CONCLUSION

This phase II single-arm trial evaluated SMES+ABX in recurrent malignant glioma using a Simon two-stage design (N=16). The primary endpoint was 4-m PFS rate, with secondary endpoints encompassing ORR, DCR, OS, NANO scale, and EORTC QLQ-C30. Safety assessments included TRAEs and SAEs. Standardized data management and bootstrap resampling were employed to ensure robustness, providing a foundation for subsequent investigations.

## Data Availability

No datasets were generated or analysed during the current study. All relevant data from this study will be made available upon study completion.

## Acknowledgements

We extend our sincere gratitude to the physicians from the Departments of Acupuncture, Oncology, Radiology, and Neurosurgery at the Third Affiliated Hospital of Zhejiang Chinese Medical University, and to Professor Lifa Huang from CIXINGTECHNOlOGY (China) for their expert guidance and collaboration in study protocol development.

## Contributors

Z.J. contributed to conceptualization, investigation, formal analysis, and original draft preparation. T.J. and Y.Z. were responsible for data curation and manuscript drafting. Q.C. conducted visualization and investigation tasks. Z.D. and Q.Y. provided resources, supervision, and validation. K.Q. and L.G. performed visualization and participated in manuscript review & editing. C.M. and X.L. oversaw study design, secured funding, managed resources, and supervised manuscript review & editing. All authors approved the final version for submission.

## Funding

The National Natural Science Foundation of China (No.82474626); the Science and Technology Plan Project jointly established by the Science and Technology Department of the National Administration of Traditional Chinese Medicine and the Zhejiang Provincial Administration of Tradi-tional Chinese Medicine (No. GZY-ZJ-KJ-24021); 2024 General Scientific Research Project of Zhejiang Provincial Department of Education - Special Project for Reform of Professional Degree Graduate Training Mode (No. Y202456327); the 2024 Zhejiang University Students Science and Technology Innovation Activity Plan (New Talent Plan) (No. 2024R410C066); Zhejiang Chinese Medical University Research Project Funding (No.2022FSYYZZ10); Zhejiang Chinese Medical University Research Project Funding (No.2023FSYYZZ09); Zhejiang Chinese Medical University Research Project Funding (No.2023FSYYZQ11)

## Competing interests

None.

## Patient and public involvement

Patients and/or the public were not involved in the design, or conduct, or reporting, or dissemination plans of this research.

## Patient consent for publication

Consent obtained directly from patient(s).

## Notes

### Competing Interest Statement

The authors have declared no competing interest.

### Clinical Trial

NCT06818331

### Funding Statement

The author(s) received no specific funding for this work.

### Author Declarations

The Medical Ethics Committee of the Third Affiliated Hospital of Zhejiang Chinese Medical University

